# Plasma NT1-tau and Aβ_42_ correlate with age and cognitive function in two large Down syndrome cohorts

**DOI:** 10.1101/2023.03.10.23287109

**Authors:** Andrew M. Stern, Kathryn L. Van Pelt, Lei Liu, Amirah K. Anderson, Beth Ostaszewski, Mark Mapstone, Sid O’Bryant, Melissa E. Petersen, Bradley T. Christian, Benjamin L. Handen, Dennis J. Selkoe, Frederick Schmitt, Elizabeth Head, the Alzheimer’s Biomarker Consortium – Down Syndrome (ABC-DS) investigators

**Affiliations:** Ann Romney Center for Neurologic Diseases, Brigham and Women’s Hospital, Harvard Medical School, Boston, MA 02115; Sanders-Brown Center for Aging, Department of Neurology, University of Kentucky, Lexington, KY 40508; Department of Neurology, University of California, Irvine, Irvine, CA 92868; University of North Texas Health Science Center, Fort Worth, TX 76107; Waisman Center, University of Wisconsin-Madison, Madison, WI 53705; Department of Psychiatry, University of Pittsburgh, Pittsburgh, PA 15213; Department of Pathology and Laboratory Medicine, University of California, Irvine, Irvine, CA 92697

**Keywords:** Down syndrome, Alzheimer disease, tau, Aβ, plasma, biomarker

## Abstract

**Introduction:** People with Down syndrome (DS) often develop Alzheimer disease (AD). Here we asked whether ultrasensitive plasma immunoassays for a tau N-terminal fragment (NT1-tau) and Aβ isoforms predict cognitive impairment.

**Methods:** Plasma NT1-tau, Aβ_37_, Aβ_40_, and Aβ_42_ levels were measured in a longitudinal discovery cohort (N = 85 participants, 220 samples) and a cross-sectional validation cohort (N = 239). We developed linear models and predicted values in the validation cohort.

**Results:** Linear mixed models for NT1-tau, Aβ_42,_ and Aβ_37:42_ were significant for age, there was no main effect of time in the discovery cohort. In cross-sectional models, NT1-tau and Aβ_42_ increased with age. NT1-tau predicted DLD scores. The discovery cohort linear model for NT1-tau predicted NT1-tau levels in the validation cohort.

**Discussion:** NT1-tau correlates with age and worse cognition in DS. Further validation of NT1-tau and other plasma biomarkers of AD neuropathology in DS cohorts is important for clinical utility.

## Background

Two populations are at nearly 100% risk for developing Alzheimer disease pathology: people carrying autosomal dominant mutations in the Aβ generation pathway (*APP* and *PSEN1/2*) and people with trisomy 21. People with trisomy 21 (Down syndrome, DS) develop amyloid plaques, then neurofibrillary tangles, and then cognitive decline beginning at an earlier age than disomic people, whereas those with partial trisomy sparing the *APP* locus do not, suggesting Aβ overproduction as a root cause of AD in DS.^1,2^ The neuropathologic features of AD due to trisomy 21 are virtually identical to sporadic AD. Further, changes in CSF Aβ_42_, CSF p-tau, hippocampal volume, amyloid PET, and dementia onset mirror the order of events in sporadic AD.^3–6^ Thus, studying the biological changes that accompany and predict onset of dementia and AD neuropathology in people with trisomy 21, a common genetic variant that occurs in many different genetic backgrounds, can help not only those with DS but also the entire AD population.^7^

In the last decade, measurement of AD-relevant analytes in plasma has been enabled by advances in mass spectrometry and ultrasensitive immunoassays. Most assays measure Aβ_40_, Aβ_42_, or different phosphorylated and/or truncated forms of tau. More recently, assays for oligomeric Aβ^8^ and Aβ_37_^9^ have emerged as potentially superior to standard Aβ_42_ and Aβ_40_ assays for distinguishing AD patients from controls. Few of these assays have been tested in people with DS, a population for whom there is great need for plasma biomarkers because of the high risk of AD and greater difficulty obtaining spinal fluid. As expected due to an extra copy of *APP*, the levels of plasma and CSF Aβ_42_ are higher in people with DS than controls.^10^ Aβ levels alone perform poorly in predicting dementia onset, though those with a declining Aβ_42:40_ ratio do have a higher risk of dementia.^10–13^ Tau biomarkers have also seen limited application to DS. Tau phosphorylated at threonine-181 (pTau181) distinguished people with DS with dementia from asymptomatic participants and correlated with CSF and brain imaging biomarkers.^14,15^

The observation that most of the tau present in body fluids is truncated led to the development of the NT1-tau assay, which detects tau fragments containing both the N-terminus and the mid-region.^16^ This assay is both sensitive and specific to AD pathology. The NT1-tau assay distinguished sporadic AD from healthy and non-AD dementia controls, in contrast to plasma neurofilament light (NfL), which could not distinguish between AD and non-AD dementia.^17^ Higher baseline NT1-tau measurement in healthy elderly controls predicted future cognitive decline, neurodegeneration by volumetric MRI, and tau PET.^18^ These results suggest plasma NT1-tau is a sensitive and specific predictor of cognitive decline due to AD neuropathology. One small cross-sectional study of NT1-tau in DS found that plasma NT1-tau levels increase with age.^10^ In the current study, we sought to validate plasma NT1-tau and Aβ isoforms as predictors of cognitive decline and neurodegeneration in people with DS in two large cohorts. We first assessed the relationship of the plasma analytes with age and cognitive function in a longitudinal cohort, and then applied the model to a separate validation cohort. We found that both plasma NT1-tau and Aβ_42_ were positively associated with age and worsening cognitive function. A linear model derived from NT1-tau values in the discovery cohort predicted the NT1-tau values in the validation cohort. The results suggest applicability of plasma NT1-tau as a biomarker of AD pathology in DS and demonstrate reproducibility in clinical use across samples.

## Methods

### Participants

Our discovery cohort was a longitudinal cohort of aging individuals with DS who were analyzed from the University of Kentucky (“UKY Cohort”). As part of the longitudinal study, participants attended visits every 6-12 months to complete a clinical assessment, blood draw, and MRI brain scan. For our analysis, only the clinical assessments and blood draw data were used. Participants were eligible if they 1) had a diagnosis of DS; 2) were over 25 years old; 3) were medically stable; 4) could complete annual visits; 5) spoke English; 6) had no neurological disease other than DS; 7) and could tolerate MRI. Participants were deemed ineligible if they were medically unstable and had changed medications in the last three months, except anxiolytic use as needed for medical procedures. Research procedures were independently reviewed and approved by the University of Kentucky Institutional Review Board. Participants completed approved protocols for informed consent or assent with guardian approval. After analysis of plasma NT1-tau and Aβ samples, the Tukey method was used to exclude outliers, with a +/-2*IQR threshold. We further excluded any participant missing *a priori* covariates or clinical outcome variables. As a result, 85 participants with 220 observations were analyzed (Table 1).

**Table 1.**
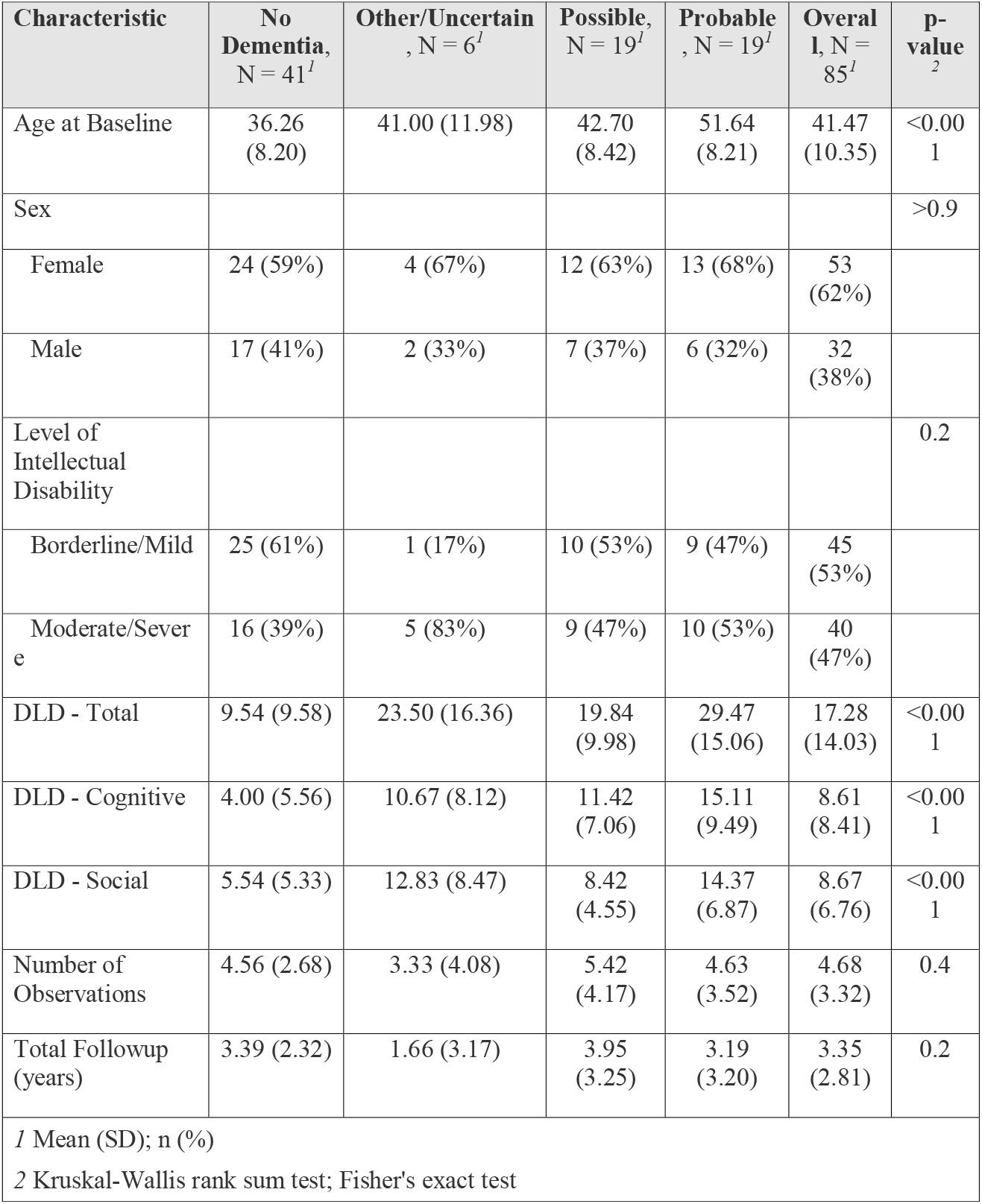
UKY Discovery Cohort characteristics.

The Alzheimer’s Biomarker Consortium – Down Syndrome (ABC-DS) served for validation. ABC-DS is a multisite cohort with ongoing enrollment.^19^ Our validation study consisted of 239 individuals with DS at their initial visit, which included blood draw, MRI, and neurocognitive assessments (Table 2). ABC-DS is conducted under IRB approved protocols, with participants and/or caregivers providing written informed consent to participate.

**Table 2.** Comparison of UKY and ABC-DS cohort demographics and plasma biomarker levels.

| Characteristic | ABCDS N = 239<sup>1</sup> | UKY, N = 85<sup>1</sup> | Overall, N = 324<sup>1</sup> | p-value<sup>2</sup> |
| --- | --- | --- | --- | --- |
| Age | 44.58 (9.74) | 41.47 (10.35) | 43.76 (9.98) | 0.016 |
| Sex |  |  |  | 0.017 |
| Female | 113 (47%) | 53 (62%) | 166 (51%) |  |
| Male | 126 (53%) | 32 (38%) | 158 (49%) |  |
| Level of Intellectual Disability |  |  |  | 0.9 |
| Borderline/Mild | 129 (54%) | 45 (53%) | 174 (54%) |  |
| Moderate/Severe | 110 (46%) | 40 (47%) | 150 (46%) |  |
| Consensus Diagnosis |  |  |  | 0.3 |
| No Dementia | 169 (71%) | 51 (60%) | 220 (68%) |  |
| Other/Uncertain | 16 (6.7%) | 8 (9.4%) | 24 (7.4%) |  |
| Possible | 34 (14%) | 15 (18%) | 49 (15%) |  |
| Probable | 20 (8.4%) | 11 (13%) | 31 (9.6%) |  |
| DLD - Total | 13.13 (14.12) | 17.28 (14.03) | 14.22 (14.19) | 0.007 |
| DLD - Cognitive | 6.77 (8.59) | 8.61 (8.41) | 7.25 (8.56) | 0.047 |
| DLD - Social | 6.36 (6.82) | 8.67 (6.76) | 6.97 (6.87) | 0.001 |
| Plasma NT1-tau (pg/ml) | 4.66 (2.05) | 3.13 (1.56) | 4.26 (2.04) | <0.001 |
| Plasma Aβ37 (pg/ml) | 45.73 (11.51) | 47.21 (17.30) | 46.12 (13.26) | >0.9 |
| Plasma Aβ40 (pg/ml) | 955.39 (454.30) | 1,620.12 (981.71) | 1,129.78 (698.92) | <0.001 |
| Plasma Aβ42 (pg/ml) | 77.23 (17.48) | 87.67 (20.51) | 79.97 (18.86) | <0.001 |
| Plasma Aβ42:Aβ40 | 0.09 (0.04) | 0.07 (0.03) | 0.09 (0.04) | <0.001 |
| Plasma Aβ37:Aβ42 | 0.62 (0.21) | 0.54 (0.15) | 0.60 (0.20) | <0.001 |
| Plasma Aβ37:Aβ40 | 0.06 (0.03) | 0.03 (0.01) | 0.05 (0.02) | <0.001 |
| <sup>1</sup>Mean (SD); n (%) |  |  |  |  |
| <sup>2</sup>Wilcoxon rank sum test; Pearson's Chi-squared test |  |  |  |  |

### Plasma handling and storage

The UKY cohort plasma was obtained under fasting conditions initially (visit 1) but transitioned to non-fasting conditions to reduce participant burden. Blood was collected into a 10 ml EDTA tube, centrifuged at 771 *g* and aliquoted into 0.5-ml volumes and frozen at -80LJ locally at the University of Kentucky. ABC-DS blood collection and processing methods were harmonized across all eight ABC-DS clinical sites. Blood was collected into a 10ml EDTA tube, then centrifuged for 10 minutes at 2,000 *g* at 4°C. The plasma fraction was aliquoted in 0.25-ml units to individual 0.5-ml siliconized cryovials and stored at -80°C at local ABC-DS clinical performance sites. The vials were shipped from the local ABC-DS sites on dry ice via overnight courier to the National Cell Repository for Alzheimer’s Disease (NCRAD) at Indiana University, where they were stored at -80°C until sent in one shipment to the Ann Romney Center for Neurologic Diseases at Brigham and Women’s Hospital in Boston, MA for analysis. The time in storage from blood draw to analysis for all ABC-DS samples was less than three years.

Upon receipt at the laboratory running the assays, aliquots of plasma were thawed, re-aliquoted, and frozen in volumes suitable for running each assay, to prevent repetitive freeze-thaw.

### Assessments

At each study visit, informants of participants in the UKY and ABC-DS cohorts completed the Dementia Scale for People with Learning Disabilities (DLD).^20^ Participants completed other behavioral and cognitive assessments in both studies, but the DLD was the only assessment common to both cohorts and was used as an independent measure in our analysis.

Informants were caregivers and/or legal guardians who were responsible for the daily care of the participants either in the home or an assisted living facility. The DLD is a 50-item informant questionnaire measuring behavioral and cognitive dysfunction. The DLD results in the following scores: sum of cognitive scores (SCS), including short-term memory, long-term memory, and spatial/temporal orientation; the sum of social scores (SOS), including speech, practical skills, mood, activity/interest, and behavioral disturbance; and a total score consisting of the combined SCS and SOS. Higher scores on the DLD indicate more deterioration.

In addition to the DLD, both the UKY and ABC-DS protocols included additional behavioral and cognitive measures that were considered for clinical diagnosis classification.^19^ Clinical diagnosis in the UKY cohort was based on NINCDS-ADRDA criteria^21^ and NACC Form D1.^22^ The consensus diagnosis was determined through expert review by a neurologist, one neuropsychologist, and either one additional neuropsychologist or one psychologist. The consensus classified participants as “no dementia,” “MCI/possible dementia,” “Alzheimer disease/probable dementia,” or “other/uncertain.” In the ABC-DS cohort, clinical diagnosis was determined by clinical consensus conference in accordance with the recommendations of the AAMR-IASSID Working Group for the Establishment of Criteria for the Diagnosis of Dementia in Individuals with Developmental Disability and was based on all available medical and cognitive testing data in reference to baseline IQ and any recent major life transitions or events.^19^ Clinical consensus decisions were made by a team of expert ABC-DS clinicians with experience in dementia and Down syndrome. ABC-DS participants received a diagnosis of cognitively stable (DS-CS), mild cognitive impairment (DS-MCI), Alzheimer’s disease dementia (DS-dementia) or were classified as “unable to determine.” Participants were classified as cognitively stable (DS-CS) if they were without cognitive or functional decline, beyond what would be expected with adult aging *per se*. Participants were classified as having MCI (DS-MCI) if they demonstrated some cognitive and/or functional decline over and above what would be expected with aging *per se*, but not severe enough to indicate the presence of dementia. Participants were categorized as having dementia (DS-dementia) if there was evidence of substantial progressive decline in cognitive function and daily living skills. An “unable to determine” category was used to characterize participants with cognitive and functional impairment that could be better explained by significant life circumstance (e.g., staff changes, family death) or conditions unrelated to AD (e.g., severe sensory loss, psychiatric diagnosis, seizure disorder). Although slightly different criteria were used between the two cohorts, we equated the UKY and ABC-DS diagnostic categories in the interest of harmonizing the data. ABC-DS “DS-CS” subjects were considered equivalent to “no dementia,” “DS-MCI” equivalent to “possible dementia,” “DS-dementia” equivalent to “probable dementia,” and “unable to determine” equivalent to “other/uncertain.”

### Plasma NT1-tau assay

Consumables and reagents other than antibodies were obtained from Quanterix (Lexington, MA). The Simoa HD-1 analyzer was used for the UKY cohort, and the Simoa HD-X analyzer was used for the ABC-DS cohort. The tau capture antibody BT2 (194–198) was conjugated onto paramagnetic beads at 1.8□mg/mL. Detector antibody Tau12 (Sigma-Aldrich) was biotinylated according to the manufacturer using a ratio of 40 parts biotinylation reagent to 1-part antibody. Plasmas were centrifuged at 14,000□×□*g* for 4□min and then diluted 1:4 with Tau 2.0 sample diluent reagent. The Tau210 standard (see below) was diluted linearly with Tau 2.0 sample diluent to a concentration range of 90–0.009□pg/mL on the HD-1 or 270-0.02 on the HD-X. Samples, standards and blanks were prepared in 1.5□mL low-binding Eppendorf tubes and were analyzed in triplicate.

The NT1-tau assay used a 3-step protocol and was performed at ambient temperature on a Simoa HD-X analyzer (Quanterix Corp.). In step 1, 100□μl of standard, blank, or sample was added to beads coated with capture antibody and mixed for 30□min. The beads were then collected and washed with wash buffer. In step 2, biotinylated detection antibody (0.6□μg/ml) was added and incubated for 10□min 30□s, and the beads were then washed three times. In step 3, 150□pM streptavidin-β-galactosidase was added, and following a further wash step, enzyme substrate (resorufin β-D-galactopyranoside) was added. The bead-bearing complexes were then resuspended and loaded into Simoa arrays, each containing 216,000 femtoliter-sized wells. The average enzyme unit per bead (AEB) was determined as described previously.^23^ Standard curves of AEB vs. Tau210 concentration were fitted to a four-parameter logistic function with 1/Y2 weighting.

### Tau210 standard

The NT1-tau assay used an in-house Tau210 untagged standard produced recombinantly in a method modified from.^24^ cDNA encoding tau protein 1-210 was cloned into pTXB1 plasmid (New England Bio) and expressed in *E. coli* BL21(DE3) using 0.4 mM IPTG for 2h starting at an OD600 of 0.6-1.0. The cells were pelleted at 3,000 *g* for 15 min and resuspended in extraction buffer (20 mM histidine, pH 6.0, 50 mM NaCl, 1 mM EDTA, 5 mM DTT, 0.1 mM PMSF, and 5% (v/v) glycerol) and snap-frozen in liquid nitrogen. Large-scale purification was performed at 4°C.

The frozen cell suspension was quickly thawed and sonicated for 6 × 30 sec with a 30-sec interval between each pulse sonication using a probe sonicator at 50 W. The homogenate was clarified by centrifugation at 21,000 *g* for 15 min. The supernatant was precipitated by incubation with 1% streptomycin sulfate (TCI) for 5 min on ice, followed by centrifugation as above. All lysates were filtered through a 0.45 mm membrane filter (Millipore). The filtrate was loaded onto a 20 ml HiPrep Q HP 16/10 column equilibrated in extraction buffer and washed extensively with ∼60 ml of the same buffer at a flow rate of 5 ml/ min until the optical density at 280 nm reached a stable baseline. Tau protein was collected into 3 ml fractions using 2 gradients followed by one isocratic elution step [gradient 1: 0.05-0.15 M NaCl (20 ml); gradient 2: 0.15-0.4 M NaCl (60 ml); isocratic: 1 M NaCl (40 ml); all in extraction buffer]. Collected fractions were screened by SDS-PAGE, and Tau-containing fractions were pooled and concentrated using a spin filter (4 kDa MWCO, Millipore). Up to 0.35 ml of the pooled-concentrated fractions was injected onto a Superdex 75 Increase 10/300 column (Cytiva) running in 50 mM ammonium acetate, pH 8.5 at a flow rate of 0.5 ml/min. 0.5-mL fractions were collected and screened by SDS-PAGE. Recombinant Tau-containing fractions were pooled and lyophilized. Columns were calibrated with Gel Filtration Standard (Bio-Rad), which ranges from 1350 to 670,000 Da.

### Plasma Aβ assays

The Aβ assays comprised Singulex sandwich ELISAs of a plate-fixed monoclonal antibody against a proprietary protein tag (Tag 1) spotted in planar arrays in 96-well microtiter plates (Planar Array Homebrew Plate, Cat. 197-0461, Quanterix). Tag 1 peptide was conjugated to the capture antibody using a bifunctional SMCC-Sulfo crosslinker: D2A6H (Cell Signaling Technologies) for Aβ_37_, QA18A67 (BioLegend) for Aβ_40_, and 1-11-3 (BioLegend) for Aβ_42_. The microtiter plates were incubated with the Tag 1-conjugated capture antibody for 30 min at RT with shaking at 525 rpm. Then, 25 μl/well of the sample were incubated for 2 hr at RT with shaking at 525 rpm. Then, 50 μl/well of biotinylated N-terminal Aβ detection antibody solution 82E1 (IBL) and HRP-streptavidin solution were both incubated for 30 min at RT with shaking at 525 rpm. Wells were washed with wash buffer between incubations. Luminol solution (ELISAbright) as the chemiluminescent substrate was added as the final step for imaging on the Quanterix Simoa SP-X system. The limit of detection (LoD) and lower limit of quantification (LLoQ) were calculated by Simoa SP-X software. All Aβ monomer peptide standards were purchased from Anaspec, dissolved in DMSO, and aliquoted.

### Statistical analysis

Participant demographics in each cohort were compared across harmonized consensus diagnoses. Kruskal-Wallis rank-sum test and Fisher’s exact test were used to compare continuous and categorical baseline participant characteristics. For all analyses, baseline age, sex, level of intellectual disability, and consensus diagnosis were selected as covariates *a priori*. To rule out other potential covariates, we also evaluated other standard blood measures of organ function as potential covariates.^25,26^ Using the first baseline visit for each available UKY participant, we evaluated the correlation between creatinine, glucose, sodium, potassium, aspartate aminotransferase, alanine aminotransferase, alkaline phosphatase, total bilirubin, hemoglobin, and platelet count. Moreover, we evaluated whether there was any effect of time of year (*i*.*e*., month) or fasting status on NT1-tau levels. There were no significant relationships between these blood tests, time of year, or fasting, and therefore they were not included in models as additional covariates (Table S1).

Because of the longitudinal nature of the UKY cohort, we first used linear mixed models to assess the change of biomarkers over time, treating the time starting from the participant’s baseline assessment as a distinct variable from baseline age. A separate linear model was generated for NT1-tau, Aβ_40_, Aβ_42_, Aβ_37_, and the ratios Aβ_37_:_40_, Aβ_37: 42_, and Aβ_40:42_. For each model, the random effects were specified using the restricted likelihood method. Random intercept versus random intercept and slope models were compared, and the random intercept and slope model was selected as there were no issues with convergence. Then main effects and interactions of the fixed effects (*i*.*e*., time and sex) were fitted using the maximum likelihood method. Model fit was selected based on loglikelihood of nested models, Akaike Information Criteria (AIC), and Bayesian Information Criteria (BIC). Analyses were completed using R packages “lme4”^27^ and “lmerTest”.^28^ As discussed below under Results, we did not observe any significant effect of time in this longitudinal analysis. Thus, linear regression models were subsequently used to measure the cross-sectional relationship of plasma biomarkers and age at baseline. Post-hoc comparisons used the Tukey correction for multiple comparisons for consensus diagnosis or intellectual disability when significant. In this cross-sectional analysis, linear regression was used to quantify the clinical associations between NT1-tau or the Aβ levels and ratios with DLD (cognitive, social, and total scores), while multinomial regression quantified the clinical associations between blood biomarker levels and consensus diagnosis. The effect of adding blood biomarkers to models was assessed by comparing model fit *via* loglikelihood, AIC, BIC and R^2^.

A total of 297 cross-sectional samples were received from ABC-DS. A Tukey outlier test was used to exclude outliers from all biomarker values, resulting in the exclusion of 58 samples, for a total of 239 in the final sample. The linear regression and multinominal regressions developed using the UKY cohort were evaluated in this ABC-DS cohort. Model fit was evaluated by comparing the predicted versus actual values along with Root Mean Squared Error (RMSE) and R^2^ retaining the model beta weights. All analyses were completed in R v4.0.3. All statistical tests were two-tailed, and the alpha-level was set at 0.05.

## Results

A total of 104 participants enrolled in the UKY longitudinal study and were included in the current analysis, with a total of 416 plasma samples. Of these, 85 participants with 220 plasma samples had sufficient demographic, clinical, and NT1-tau and Aβ plasma values for analysis (Table 1).

Except for NT1-tau, Aβ_42,_ and Aβ_37:42_, no linear mixed model had significantly better model fit than the null model (all p > 0.05, for full results see Table S2). For NT1-tau and Aβ_37:42_, there was no significant main effect of time (Fig S1A, C) while there was a significant but unconvincing main effect of time for Aβ_42_ (p = 0.03) (Fig S1B). However, examining spaghetti plots of these plasma measures showed a visual increase with age for NT1-tau, particularly after the age of 40 years (Fig S1D). Therefore, the longitudinal nature of the UKY cohort did not improve the predictive properties of the biomarkers due to a strong age effect, and this cohort was then treated as cross-sectional. The baseline visit from each of the 85 UKY participants was used in a cross-sectional analysis to quantify the relationship between baseline age, sex, level of intellectual disability, consensus diagnosis and each blood biomarker. Across their final consensus diagnosis, there were significant differences in age, DLD - Total, DLD - Cognitive, and DLD - Social scores (Table 1). These differences were expected given the known relationship between age, cognitive function, and consensus diagnosis. The NT1-tau linear regression model was significant (F(6, 78) = 4.98; p < 0.001; R^2^_adj_ = 0.22; RMSE = 1.32). Age was significantly positively associated with NT1-tau, whereby every one-year increase in age was associated with a 0.05 pg/ml increase in NT1-tau (95% CI: 0.02 - 2.93, t = 2.93, p = 0.004) (Figure 1A). Full model results are available in Table S3. There were no other significant main effects observed (sex, level of ID, consensus diagnosis all p > 0.05).

**Fig 1.**
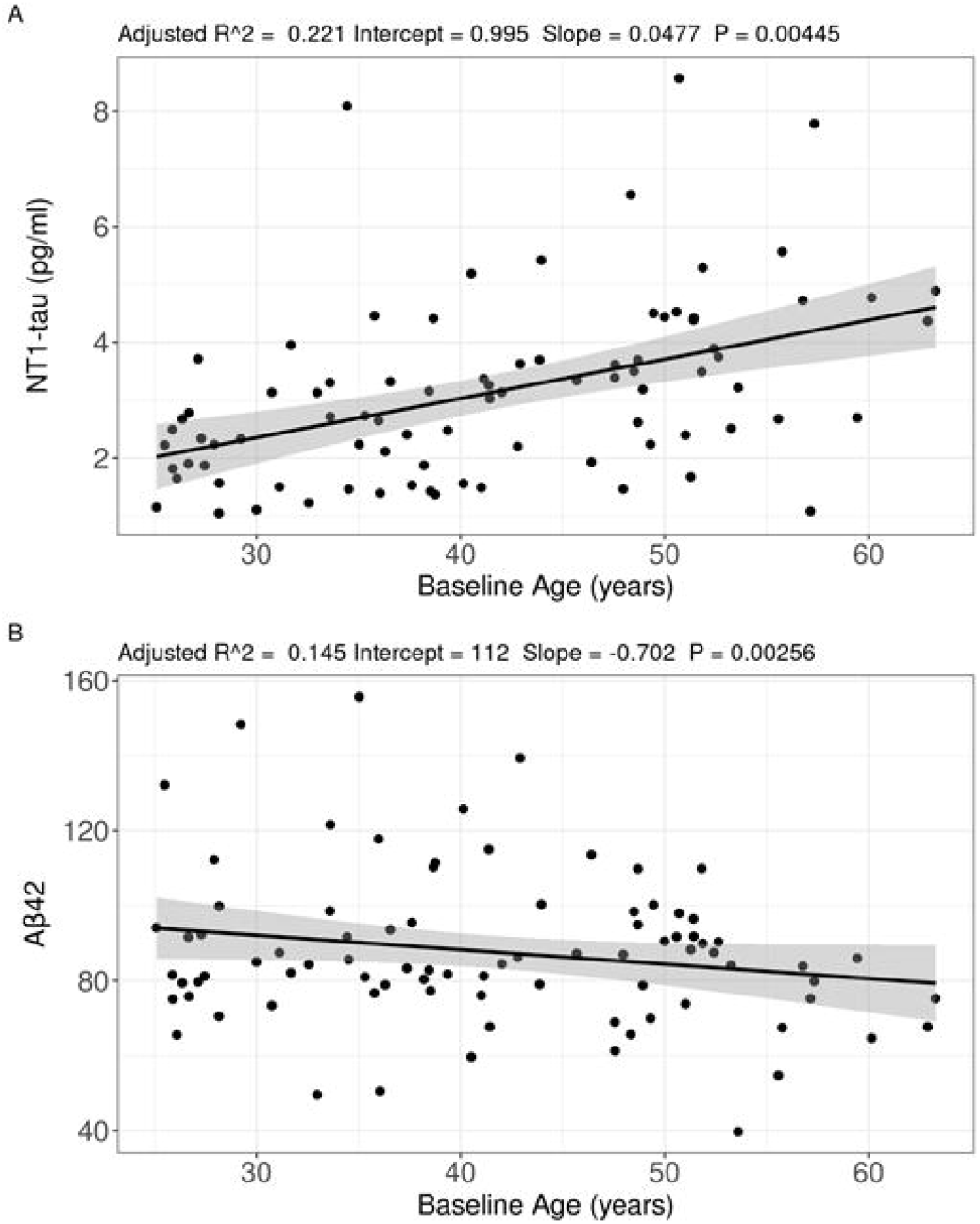
Cross-sectional linear regression models in the discovery cohort. Correlation of (A) NT1-tau and (B) Aβ_42_ with baseline age. Covariates included sex, level of intellectual disability, and consensus diagnosis.

For Aβ_42_, the linear regression model was significant (F(6, 78) = 3.37; p = 0.005; R^2^_adj_ = 0.14; RMSE = 18.18) (Figure 1B). Age was significantly associated with Aβ_42_, whereby every one-year increase in age was associated with a 0.70 pg/ml decrease in Aβ_42_ (95% CI: -1.15, -0.25, t = -3.12, p = 0.003). After using Tukey correction for multiple comparisons, “probable dementia” had higher Aβ_42_ than the no dementia group. Full model results are available in Table S4. The following models were not significant: Aβ_40_ (F(6, 78) = 0.76; p = 0.606), Aβ_37_ (F(6, 78) = 0.75; p = 0.613), Aβ_42:40_, (F(6, 78) = 1.05; p = 0.400), Aβ_37:42_ (F(6, 78) = 1.26; p = 0.287), Aβ_37:40_ (F(6, 78) = 0.64; p = 0.695).

When examining the relationship between NT1-tau and DLD, NT1-tau was not associated with DLD - cognitive, DLD - Social, or DLD - Total when age, sex, level of intellectual disability, and diagnosis were included in the model (both p > 0.05). However, NT1- tau was significantly positively associated with higher (worse) DLD - Cognitive (β = 1.76, t = 3.08, p = 0.003), DLD - Social (β = 1.44, t = 3.16, p = 0.002), and DLD - Total (β = 1.20, t = 3.41, p = 0.001) scores when Aβ_40_, Aβ_42_, and Aβ_37_ were the only covariates (Figure 2). Aβ_40_, Aβ_42_, and Aβ_37_ were not significantly associated with any DLD scores when all were included in a model with NT1-tau (all p > 0.05).

**Fig 2.**
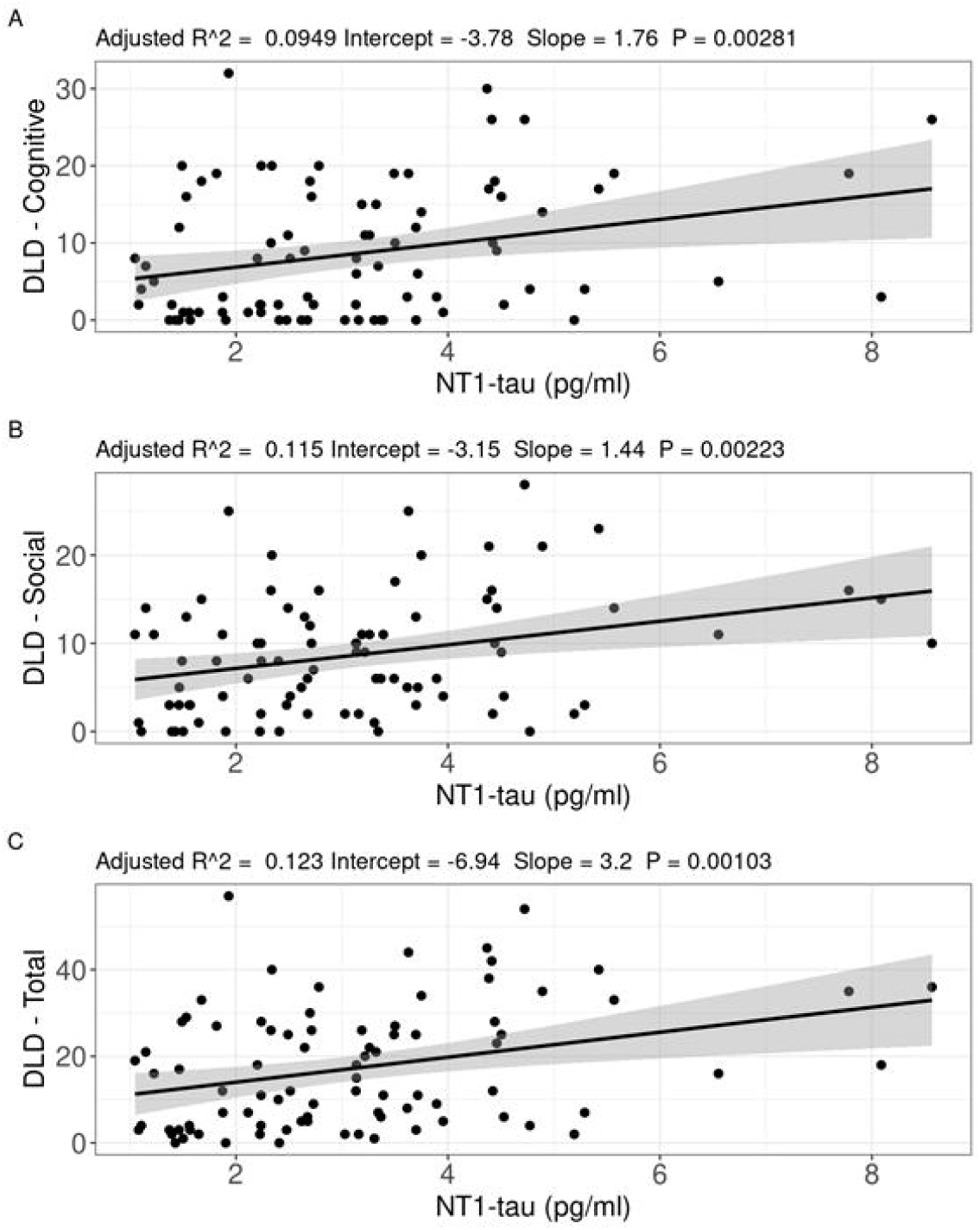
Plasma NT1-tau correlates with cognitive impairment. Correlation of plasma NT1-tau with (A) DLD – Cognitive, (B) DLD – Social, and (C) DLD – Total in the cross-sectional analysis of the discovery cohort. Covariates included plasma Aβ_40_, Aβ_42_, and Aβ_37._ Significant effects were lost when age was included as a covariate (see text).

To test the external validity of the models developed on the UKY discovery cohort, the linear regression models for each biomarker were tested on the independent ABC-DS validation cohort. Linear regressions developed from the UKY cohort were applied to the ABC-DS cohort to predict the levels of NT1-tau, Aβ_42,_ Aβ_40_, Aβ_37_, Aβ_37:40_, Aβ_37:42_, and Aβ_40:42_. There was a significant correlation between predicted and actual NT1-tau values in the ABC-DS cohort using the UKY model (r = 0.38; p < 0.001; Figure 3A). While the Aβ_42_ linear regression model was statistically significant in the UKY cohort, it did not predict the levels of Aβ_42_ in the validation ABC-DS cohort (r = 0.01, p = 0.85) (Figure 3B).

**Fig 3.**
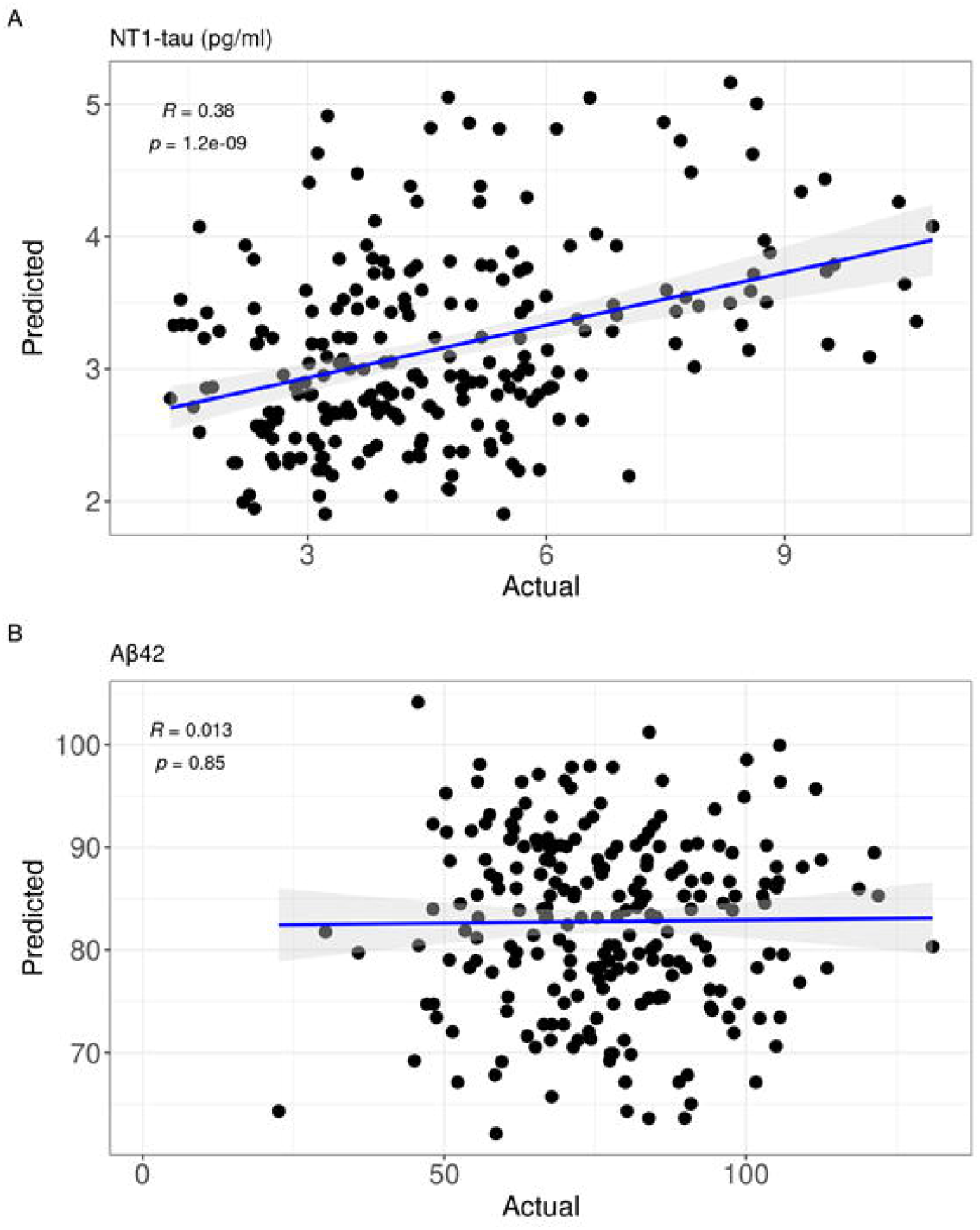
NT1-tau linear regression model in discovery cohort predicts plasma NT1-tau in validation cohort. The statistically significant linear models for NT1-tau (A) and Aβ_42_ (B) in the discovery cohort were used to generate predicted values in the validation cohort, which were then correlated with the actual results. NT1-tau but not Aβ_42_ produced a significant correlation.

## Discussion

Here we sought to characterize changes in plasma NT1-tau and Aβ isoform levels with respect to time, age, diagnosis, and cognitive impairment in people with DS. We started with the longitudinal UKY cohort, finding in a linear mixed model that age predicted NT1-tau levels. As seen by others for plasma neurofilament light and for CSF biomarkers,^29^ there appeared to be a stereotypical inflection point around age 40 at which NT1-tau accelerated upwards, which was visible using spaghetti plots (Fig S1D). A similar inflection pattern was not observed in plasma Aβ. Because of variability in longitudinal measurements of NT1-tau and Aβ isoforms across multiple years, combined with a strong age effect, we could not detect progressive increases or decreases in these plasma biomarkers within the same individual (*i*.*e*., an effect of time in the linear mixed model). Hence, we used the baseline levels in linear regression models. In these models, NT1-tau correlated with age, and was positively correlated with higher scores on the DLD (indicating worsening dysfunction). Aβ_42_ showed the opposite, weaker, correlation.

Because of the strong effect of age on cognitive decline in the DS population, we did not see a clear improvement in prediction of cognitive decline by adding plasma biomarkers to the model. The linear models used to predict NT1-tau in the discovery cohort successfully predicted levels in the validation cohort. Overall, our results suggest that plasma NT1-tau is a valid marker of cognitive impairment in a broad DS population.

The variable measures across timepoints within individuals (Fig S1D) is important to consider for NT1-tau, but also other plasma biomarkers. In a recent study, even within carefully controlled visits by the same researchers only 6-10 weeks apart, there was 20% or more test-retest variability of plasma P-tau217, neurofilament light, and glial fibrillary acidic protein.^30^ In a person who might undergo sampling years apart by different practitioners, pre-analytical variation will vary even more than this. While the NT1-tau values from the discovery cohort predicted the values in the validation cohort (Fig. 3A), the prediction was not perfect (r = 0.380 and may demonstrate the difficulties in applying academic cohort models to predict disease in real, individual, people. In the future, we hope to refine the longitudinal stability of the NT1-tau and other plasma biomarker assays to improve real-world applicability.

## Conclusions

Plasma NT1-tau is a biomarker of cognitive impairment in people with DS, moreso than Aβ isoforms. Longitudinal analytical variability and strong age effects in the DS population may impact the use of plasma biomarkers.

## Data Availability

All data produced in the present study are available upon reasonable request to the authors.

## Acknowledgements

The authors would like to thank the patients and families who contributed their time and plasma to this study.

^#^Alzheimer’s Biomarker Consortium-Down Syndrome (ABC-DS) Investigators: Howard J. Aizenstein, MD PhD; Beau M. Ances, MD PhD; Howard F. Andrews, PhD; Karen Bell, MD; Rasmus M. Birn, PhD; Adam M. Brickman, PhD; Peter Bulova, MD; Amrita Cheema, PhD; Kewei Chen, PhD; Bradley T. Christian, PhD; Isabel Clare, PhD; Lorraine Clark, PhD; Ann D. Cohen, PhD; John N. Constantino, MD; Eric W. Doran, MS; Anne Fagan, PhD; Eleanor Feingold, PhD; Tatiana M. Foroud, PhD; Benjamin L. Handen, PhD; Sigan L. Hartley, PhD; Elizabeth Head, PhD; Rachel Henson, PhD; Christy Hom, PhD; Lawrence Honig, MD; Milos D. Ikonomovic, MD; Sterling C Johnson, PhD; Courtney Jordan, RN; M. Ilyas Kamboh, PhD; David Keator, PhD; William E. Klunk, MD PhD; Julia K. Kofler, MD; William Charles Kreisl, MD; Sharon J. Krinsky-McHale, PhD; Florence Lai, MD; Patrick Lao, PhD; Charles Laymon, PhD; Joseph H Lee, PhD; Ira T. Lott, MD; Victoria Lupson, PhD; Mark Mapstone, PhD; Chester

A. Mathis, PhD; Davneet Singh Minhas, PhD; Neelesh Nadkarni, MD; Sid O’Bryant, PhD; Deborah Pang, MPH; Melissa Petersen, PhD; Julie C. Price, PhD; Margaret Pulsifer, PhD; Michael Rafii MD PhD; Eric Reiman, MD; Batool Rizvi, MS; Herminia Diana Rosas, MD; Nicole Schupf, PhD; Wayne P. Silverman, PhD; Dana L. Tudorascu, PhD; Rameshwari Tumuluru, MD; Benjamin Tycko, MD PhD; Badri Varadarajan, PhD; Desiree A. White, PhD; Michael A. Yassa, PhD; Shahid Zaman, MD PhD; Fan Zhang, PhD

## Consent Statement

All participants (or their guardians when appropriate) provided written informed consent under Institutional Review Board approval for this study.

