## Supplementary Information for "Plasma NT1-tau and Aβ_42_ correlate with age and cognitive function in two large Down syndrome cohorts"

**Supplementary Tables**

| **Covariate** | **Test Statistic** | **p-value** |
| --- | --- | --- |
| Creatine | Pearson Correlation | p = 0.062 |
| Month (time of year) | Kruiskal-Wallis | p = 0.370 |
| Fasting | Kruiskal-Wallis | p = 0.075 |
| Glucose | Pearson Correlation | p = 0.380 |
| Sodium | Pearson Correlation | p = 0.470 |
| Potassium | Pearson Correlation | p = 0.610 |
| AST | Pearson Correlation | p = 0.510 |
| Alanine Aminotransferase | Pearson Correlation | p = 0.21 |
| Alkaline Phosphatase | Pearson Correlation | p = 0.99 |
| Bilirubin | Pearson Correlation | p = 0.13 |
| Hemaglobin | Pearson Correlation | p = 0.98 |
| Platelet | Pearson Correlation | p = 0.067 |

Table S1. Effect of blood chemistries on plasma NT1-tau levels.

| NT1-tau | | | | | | | |  |
| --- | --- | --- | --- | --- | --- | --- | --- | --- |
|  |  | Sum Sq | Mean Sq | NumDF | DenDF | F value | Pr(>F) | Model Fit |
|  | Baseline Age | 10.0905 | 10.0905 | 1 | 81.58 | 12.6963 | 0.0006154 | X2(7) = 34.90; p < 0.001 |
|  | Sex | 0.3061 | 0.3061 | 1 | 67.045 | 0.3852 | 0.5369484 |  |
|  | ID Level | 0.0011 | 0.0011 | 1 | 67.531 | 0.0014 | 0.9700154 |  |
|  | Time | 1.8142 | 1.8142 | 1 | 23.429 | 2.2827 | 0.1441947 |  |
|  | Final Diagnosis | 2.7582 | 0.9194 | 3 | 73.485 | 1.1568 | 0.3321128 |  |
| Aβ42 | | | | | | | |  |
|  |  | Sum Sq | Mean Sq | NumDF | DenDF | F value | Pr(>F) | Model Fit |
|  | Baseline Age | 611.65 | 611.65 | 1 | 77.362 | 3.2284 | 0.07628 | X2(9) = 17.58; p = 0.040 |
|  | Sex | 889.44 | 889.44 | 1 | 66.547 | 4.6946 | 0.03384 |  |
|  | ID Level | 55.14 | 55.14 | 1 | 66.671 | 0.2911 | 0.59134 |  |
|  | Time | 1012.26 | 1012.26 | 1 | 17.111 | 5.3428 | 0.03352 |  |
|  | Final Diagnosis | 428 | 142.67 | 3 | 73.961 | 0.753 | 0.52409 |  |
| Aβ40 | | | | | | | |  |
|  |  | Sum Sq | Mean Sq | NumDF | DenDF | F value | Pr(>F) | Model Fit |
|  | Baseline Age | 21551 | 21551 | 1 | 82.724 | 0.3026 | 0.58372 | X2(9) = 12.35; p = 0.195 |
|  | Sex | 1325 | 1325 | 1 | 82.48 | 0.0186 | 0.89185 |  |
|  | ID Level | 99145 | 99145 | 1 | 82.678 | 1.3922 | 0.24141 |  |
|  | Time | 489586 | 489586 | 1 | 29.687 | 6.875 | 0.01365 |  |
|  | Final Diagnosis | 131872 | 43957 | 3 | 82.763 | 0.6173 | 0.60575 |  |
| Aβ37 | | | | | | | |  |
|  |  | Sum Sq | Mean Sq | NumDF | DenDF | F value | Pr(>F) | Model Fit |
|  | Baseline Age | 35.57 | 35.57 | 1 | 74.754 | 0.3896 | 0.53442 | Singular Fit - X2(9) = 9.72; p = 0.374 |
|  | Sex | 34.38 | 34.38 | 1 | 68.233 | 0.3765 | 0.54155 |  |
|  | ID Level | 278.32 | 278.32 | 1 | 68.452 | 3.0479 | 0.08533 |  |
|  | Time | 489.21 | 489.21 | 1 | 32.894 | 5.3574 | 0.02702 |  |
|  | Final Diagnosis | 142.77 | 47.59 | 3 | 72.584 | 0.5212 | 0.66906 |  |
| Aβ42:Aβ40 | | | | | | | |  |
|  |  | Sum Sq | Mean Sq | NumDF | DenDF | F value | Pr(>F) | Model Fit |
|  | Baseline Age | 0.00000039 | 3.88E-07 | 1 | 86.331 | 0.004 | 0.94954 | X2(9) = 11.41; p = 0.249 |
|  | Sex | 0.00000129 | 1.29E-06 | 1 | 67.209 | 0.0134 | 0.9082 |  |
|  | ID Level | 0.00000693 | 6.93E-06 | 1 | 66.885 | 0.0718 | 0.78949 |  |
|  | Time | 0.00018668 | 1.87E-04 | 1 | 14.835 | 1.9367 | 0.18454 |  |
|  | Final Diagnosis | 0.00080706 | 2.69E-04 | 3 | 71.307 | 2.791 | 0.04661 |  |
| Aβ37:Aβ42 | | | | | | | |  |
|  |  | Sum Sq | Mean Sq | NumDF | DenDF | F value | Pr(>F) | Model Fit |
|  | Baseline Age | 0.044871 | 0.044871 | 1 | 70.475 | 5.8928 | 0.01776 | Singular Fit - X2(9) = 20.68; p = 0.014 |
|  | Sex | 0.057632 | 0.057632 | 1 | 38.652 | 7.5687 | 0.008991 |  |
|  | ID Level | 0.044245 | 0.044245 | 1 | 37.997 | 5.8107 | 0.020875 |  |
|  | Time | 0.010995 | 0.010995 | 1 | 97.568 | 1.444 | 0.232399 |  |
|  | Final Diagnosis | 0.076387 | 0.025462 | 3 | 42.643 | 3.3439 | 0.027836 |  |
| Aβ37:Aβ40 | | | | | | | |  |
|  |  | Sum Sq | Mean Sq | NumDF | DenDF | F value | Pr(>F) | Model Fit |
|  | Baseline Age | 8.49E-05 | 8.49E-05 | 1 | 88.071 | 2.7593 | 0.1002 | Singular Fit - X2(9) = 10.91; p = 0.282 |
|  | Sex | 7.33E-05 | 7.33E-05 | 1 | 81.047 | 2.3818 | 0.1267 |  |
|  | ID Level | 2.19E-05 | 2.19E-05 | 1 | 81.254 | 0.7127 | 0.401 |  |
|  | Time | 2.58E-05 | 2.58E-05 | 1 | 108.841 | 0.8394 | 0.3616 |  |
|  | Final Diagnosis | 9.48E-05 | 3.16E-05 | 3 | 84.313 | 1.0274 | 0.3847 |  |

Table S2. Full linear mixed model effects.

| NT1-tau | | | | |
| --- | --- | --- | --- | --- |
|  | **Estimate** | **Std. Error** | **t value** | **Pr(>\|t\|)** |
| (Intercept) | 0.995 | 0.676 | 1.471 | 0.145 |
| Age | 0.048 | 0.016 | 2.93 | 0.004 |
| Sex - Male | −0.290 | 0.311 | −0.933 | 0.354 |
| ID Level - Moderate/Severe | −0.041 | 0.308 | −0.133 | 0.894 |
| Diagnosis - Other/Uncertain | 0.211 | 0.533 | 0.395 | 0.694 |
| Diagnosis - Possible | 0.593 | 0.427 | 1.39 | 0.168 |
| Diagnosis Probable | 1.242 | 0.504 | 2.463 | 0.016 |

Table S3. NT1-tau linear regression model results.

| Aβ42 | | | | |
| --- | --- | --- | --- | --- |
|  | **Estimate** | **Std. Error** | **t value** | **Pr(>\|t\|)** |
| (Intercept) | 111.941 | 9.348 | 11.974 | 0 |
| Age | −0.702 | 0.225 | −3.117 | 0.003 |
| Sex - Male | −7.612 | 4.304 | −1.769 | 0.081 |
| ID Level - Moderate/Severe | 4.121 | 4.256 | 0.968 | 0.336 |
| Diagnosis - Other/Uncertain | 13.258 | 7.363 | 1.801 | 0.076 |
| Diagnosis - Possible | 16.169 | 5.899 | 2.741 | 0.008 |
| Diagnosis Probable | 12.931 | 6.971 | 1.855 | 0.067 |

Table S4. Aβ_42_ linear regression model results.

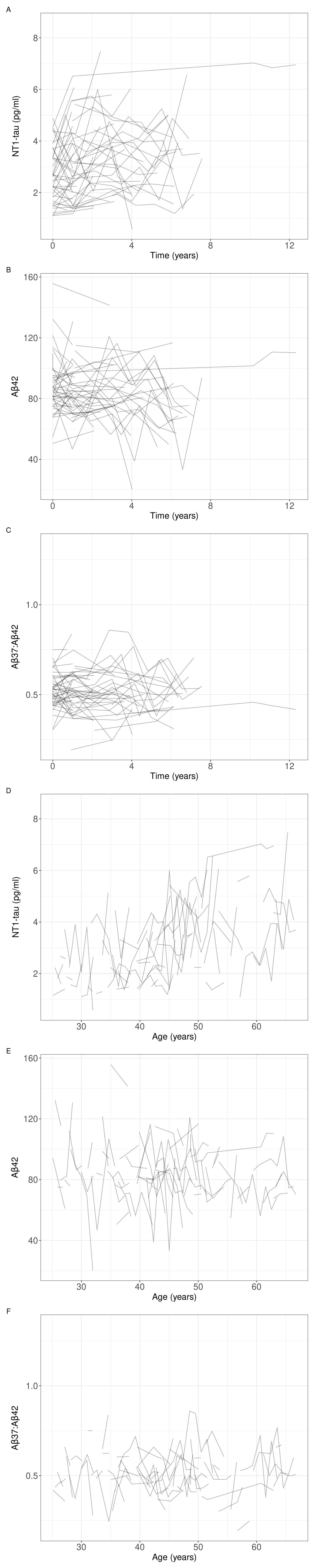

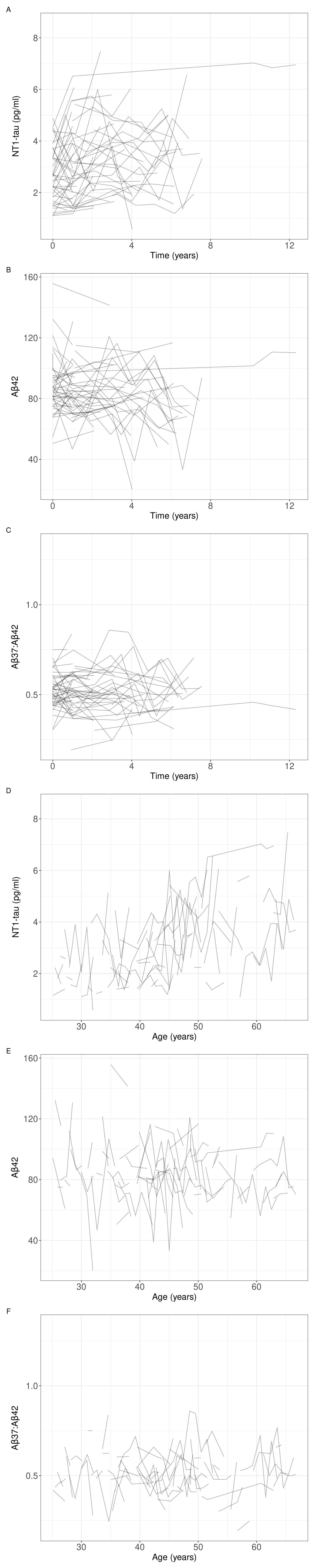

**Fig S1**. Spaghetti plots with respect to time since first blood draw (A, B, C) or age (D, E, F) of NT1-tau, Aβ42, or Aβ37:42 ratio.
